# Short-term Preeclampsia Prediction: Cutoff Variations for sFlt-1/PlGF in U.S. Patients with or without Hypertensive Disorders

**DOI:** 10.1101/2025.10.24.25337910

**Authors:** Yaxin Li, Kristen Cagino, Jim Yee, Caroline Andy, Dajana Borova, Ayush Shah, Isla Racine, Tracy B. Grossman, Zhen Zhao

**Author notes:** Corresponding authors Tracy B. Grossman, 506 6th St, Miner Pavillion - 4th floor, Brooklyn NY 11215, USA, Zhen Zhao, 525 East 68^th^ Street, F-701, New York, NY, 10065, USA. Contribute equally (Yaxin Li, Kristen Cagino).

## Abstract

**Background:** Preeclampsia (PE) is a complex disorder with significant maternal and fetal risks. The soluble fms-like tyrosine kinase-1 (sFlt-1) and placental growth factor (PlGF) ratio shows promise as a diagnostic tool, but its adoption in the U.S. is limited, due to a lack of widely available testing platforms, locally conducted studies based on the U.S. population and clear, evidence-based guidance on test implementation, including appropriate cutoff values.

**Patients/Materials and Methods:** This is a cohort study evaluating the sFlt-1/PlGF ratio for predicting PE within two weeks among pregnant individuals (≥18 years old, gestational age ≥20 weeks. Serum samples were collected during routine prenatal visits or using leftover samples from triage evaluations. Patients diagnosed with PE before sample collection, those with postpartum samples, or those missing delivery or diagnostic data were excluded. sFlt-1/PlGF ratios were measured using Roche Elecsys assays, and predictive performance was assessed by operating characteristics (ROC) curve analysis with logistic regression, bootstrapping, and cross-validation.

**Results:** The sFlt-1/PlGF ratio was significantly higher in hypertensive patients (n = 119) than in non-hypertensive patients (n = 346) (*p* < 0.001). PE developed in 9.7% of all patients and 37.0% of hypertensive patients within two weeks. ROC analysis showed an area under the curve (AUC) of 0.92 (95%CI: 0.88-0.95) for the overall population, higher than 0.82 (95%CI: 0.73-0.89) in the hypertensive group. The optimal cutoff for the overall population was 33 (95% CI: 26-47), yielding a sensitivity of 93.8% (95%CI: 85.4-100.0), specificity of 81.3% (95%CI: 76.8-85.8), negative predictive value (NPV) of 99.2% (95%: 98.0-100.0), and positive predictive value (PPV) of 35.3% (95%CI: 25.1-45.5). For the hypertensive group, the optimal cutoff was 50 (95% CI: 26-84), with a sensitivity of 81.7% (95%: 61.8-97.4), specificity of 73.1% (95%CI: 53.3-90.4), NPV of 87.6% (95%CI: 76.6-97.7), and PPV of 65.2% (95%CI: 50.0-82.5).

**Conclusions:** The sFlt-1/PlGF ratio shows high NPVs for ruling-out PE, but moderate PPVs, limiting its rule-in ability. Additionally, our findings indicate different performance in the overall and hypertensive patients, highlighting the need for further research to refine diagnostic thresholds and improve clinical guidance.

## Introduction

Preeclampsia (PE) occurs in approximately 2-7% of pregnancies depending on population, and poses a substantial risk to both mother and fetus [1], leading to an estimated 70,000 maternal and half a million fetal deaths globally each year [2]. The diagnosis of PE depends on the presence of hypertension in addition to several clinical and laboratory findings. Adding to this complexity is the fact that there are multiple overlapping clinical phenotypes that can mimic PE, such as chronic hypertension, systemic lupus erythematosus and chronic kidney disease [3–5]. Therefore, the use of a reliable serum biomarker would be helpful in predicting PE in patients with challenging clinical presentations. Having the ability to efficiently rule-in or -out PE can help to avoid prolonged hospitalization and associated healthcare costs.

Soluble fms-like tyrosine kinase-1 (sFlt-1) and placental growth factor (PlGF) are key angiogenic biomarkers that play a significant role in the pathogenesis of PE and abnormal levels of each can indicate placental dysfunction [1,6]. In normal pregnancies, sFlt-1 maintains healthy vascular function; however, in PE, increased sFlt-1 reduces vascular endothelial growth factor and PlGF levels, leading to endothelial dysfunction [3]. The sFlt-1/PlGF ratio has been proposed as an important predictor of PE [6–8], and typically increases weeks before symptoms appear. It has also been used to help determine if PE is present in patients with conditions that mimic PE, such as idiopathic thrombocytopenic purpura [9–12]. A high sFlt-1/PlGF ratio suggests the presence of PE or a higher risk of PE development, which can lead to the decision to intensify maternal and fetal surveillance, or to delivery, depending on the gestational age (GA). Incorporating this ratio in diagnostic protocols may also help to curb healthcare costs by limiting inpatient hospital admission and intensive monitoring to those at highest risk for PE development [7,13,14].

Several commercialized immunoassays, such as Triage PLGF test (Quidel), DELFIA Xpress PLGF 1-2-3 test and DELFIA Xpress sFlt-1 kit (PerkinElmer), Elecsys sFlt-1/PlGF (Roche Diagnostics) and BRAHMS PlGF and sFlt-1 KRYPTOR (Thermo Fisher Scientific), are available to measure sFlt-1 and PlGF. However, it has been reported that these assays are manufacturer-specific, and their cutoffs are not interchangeable [15–17]. Although Thermo Fisher Scientific’s BRAHMS PlGF and sFlt-1 KRYPTOR tests along with Roche Diagnostics’ Elecsys sFlt-1/PlGF ratio tests have received U.S. FDA approval in May 2023 and February 2025, respectively, their FDA-approved intended use is quite restrictive, limited to hospitalized patients with hypertensive disorders for progression to PE with severe features within two weeks of presentation. The need to explore this broader intended use in the U.S. is critical, as outpatients, who are not under constant medical surveillance, offer greater insights into PE development. Furthermore, the American College of Obstetricians and Gynecologists (ACOG) Clinical Practice Update, based on the KRYPTOR tests, cannot be generalized to other methods or assays [18]. Furthermore, there are currently no U.S. practice guidance available based on other assays, partially due to a lack of FDA cleared tests aside from the KRYPTOR tests and limited U.S.-based data. The United Kingdom’s National Institute of Health and Care Excellence (NICE) guideline is based on multiple assays [19]. However, multiple different cutoff values for each test have been suggested in the NICE guideline depending on GAs, intentions (diagnosis and prediction), decision rules (rule-in and rule-out), which can complicate the implementation.

Many studies outside of the U.S. have investigated the use of the sFlt-1/PlGF ratio for predicting PE within a certain amount of time or diagnosing PE at different GAs using the Roche Elecsys sFlt-1/PlGF ratio [20–23]. Verlohren et al. established GA-specific cutoff values (early-20 0/7 to 33 6/7 and late-34 0/7 to delivery) for the sFlt-1/PlGF ratio in the diagnosis of PE [20]. Later, the PROGNOSIS study identified a single cutoff point for the sFlt-1/PlGF ratio from GA of 24 0/7 to 36 6/7 weeks [24]. This cut-off value was shown to rule-out PE within one week and rule-in PE within four weeks in cases of suspected PE [24]. Furthermore, it has been reported that the sFlt-1/PlGF ratio can help discriminate chronic hypertension from superimposed PE--a subtype of PE that occurs in patients with preexisting chronic hypertension [25]. This distinction is challenging because significant proteinuria frequently coexists in patients with chronic hypertension due to nephrosclerosis resulting from long-standing hypertension [26]. A limited number of U.S. studies have assessed the sFlt-1/PlGF ratio in pregnancies [22,23,27,28]. The patients included in these studies were primarily those presenting acutely with clinical signs or symptoms concerning (suspected) PE. Moreover, previous U.S. studies often focused on severe PE [27,28]. However, mild PE should not be overlooked, as it can progress to severe PE or eclampsia without early and proper management. Additionally, the incidence of PE varies globally, with higher rates observed in developing countries [29]; However, the U.S. likely faces elevated risks due to higher rates of obesity, advanced maternal age, chronic conditions, and multiple gestations. Validating the biomarker in a diverse U.S. cohort is crucial, as demographic differences and obesity prevalence may influence its diagnostic performance and optimal cutoffs. It is important to note that the diagnostic criteria of PE have undergone significant changes in the U.S. over the past several years [30–32], adding complexity to the interpretation of sFlt-1/PlGF data from most of the previous U.S. studies utilizing older generations of the PE diagnostic criteria. Thus, there is a research need to investigate its clinical utility in a broader U.S. population using the updated ACOG criteria.

The aim of this study was to evaluate the clinical performance of the Roche sFlt-1/PlGF ratio in a U.S. population, with and without hypertensive disorders in predicting PE. Predicting and differentiating PE from other hypertensive disorders such as chronic hypertension and gestational hypertension is crucial because PE carries with it a higher risk of morbidity and mortality and requires more intensive assessment and intervention [28,33].

## Methods

### Participants recruitment and sample collection

This study was approved by the institutional review board (IRB) at Weill Cornell Medicine (IRB#:1806019375). A subset of pregnant individuals at or above the age of 18 (GA≥ 20 weeks) receiving prenatal care at New York-Presbyterian -Weill Cornell Hospital between December 1, 2018, and March 12, 2020, were enrolled in this study. Patients already diagnosed with PE before sample collection and those whose blood samples were collected in the postpartum period were excluded from the study. Additionally, patients with missing delivery and PE diagnosis data were also excluded. The study population comprised pregnant women whose blood samples were collected during routine prenatal care visits—with written informed consent obtained—and from patients evaluated for PE in Triage, whose leftover serum samples were used (waived consent approved by the same IRB). The residual serum samples were stored at -80°C until analysis. This study was conducted in accordance with the principles of the Declaration of Helsinki.

### Diagnosis criteria

Hypertension is defined as systolic blood pressure of 140 mmHg or higher, or diastolic blood pressure of 90 mmHg or higher, measured on at least two occasions at least four hours apart [34]. Gestational hypertension is diagnosed in patients with newly onset hypertension, occurring after 20 weeks of gestation [35]. Chronic hypertension is diagnosed when hypertension is present before 20 weeks of gestation [36]. Hypertensive disorders of pregnancy comprise chronic hypertension, gestational hypertension, PE/eclampsia [37].

The PE is defined according to the ACOG diagnostic criteria of published in 2020 [33,38]: Hypertension with at least one of the following (1) Proteinuria of 300 mg or more in a 24-hour urine specimen, or a protein/creatinine ratio of 0.3 or higher in a random urine specimen, or a dipstick reading of 2+ or higher when a quantitative measurement is unavailable; (2) A platelet count below 100,000/µL; (3) Serum creatinine levels exceeding 1.1 mg/dL, or a doubling of the creatinine concentration in the absence of other kidney diseases; (4) Liver transaminases at least twice the normal upper limit according to the local laboratory; (5) Pulmonary edema; (6) New-onset and persistent headaches unresponsive to medication; (7) Vision changes, including blurred vision; (8) Severe persistent upper right abdominal pain unresponsive to medication; (9) Severe range blood pressures (≥ 160/110 mm Hg) on two occasions at least 4 hours apart.

### Data collection

Demographic factors including patient age, race, GA at time of blood draw, first-trimester BMI, smoking status and parity were collected from electronic medical records (EMR). Development of PE was also collected from EMR by investigators practicing clinical obstetrics and those trained to assess for the diagnostic criteria for PE.

### Laboratory testing of sFlt-1 and PlGF

Levels of sFlt-1 and PlGF were quantified using Cobas e411 analyzer (Roche Diagnostics), a fully automated electrochemiluminescence immunoassay platform with detection limits of 10 pg/mL and 3 pg/mL, respectively. At the time of this study, these assays were not yet cleared for clinical use by the U.S. FDA and were for research use only.

### Statistical analysis

We performed Receiver Operating Characteristic (ROC) curve analysis to evaluate the predictive performance of the sFlt-1/PlGF ratio, which was built using logistic regression. The analysis was conducted using stratified 5-fold cross-validation, ensuring an equal distribution of outcome classes across each fold. The primary outcome was the sFlt-1/PlGF ratio’s ability to predict the development of PE within two weeks in both the overall study population and a subgroup of hypertensive (pregnant individuals present with gestational hypertension or chronic hypertension at enrollment) patients. To obtain reliable estimates of performance metrics and assess variability, we applied bootstrapping resampling (n=1000 iterations), which allowed for the calculation of 95% confidence intervals (CIs). The ROC curve was generated by plotting true positive rate (sensitivity) against false positive rate (1-specificity), and the area under the curve (AUC) was used as the primary measure of model discrimination. The optimal cutoff values were determined using the Youden index. In addition to the ROC curve and AUC, we reported other performance metrics including AUC, optimal cutoff, sensitivity, specificity, positive predictive value (PPV), and negative predictive value (NPV), F1-score, positive likelihood ratio (LR+) and negative likelihood ratio (LR-) and the corresponding 95% CIs. The *p* values comparing the performance metrics between the overall population and the hypertensive subgroup were calculated using Z-tests. Baseline characteristics between PE and non-PE cohorts were compared using the Mann-Whitney U test and Chi-squares. Statistical analyses were conducted using GraphPad Prism version 8 (GraphPad Software Inc.) or Python.

## Results

### Baseline characteristics

Blood samples from 514 pregnant women were initially included in this study. Participants diagnosed with PE prior to blood collection (n=9), those with samples collected during the postpartum period (n=25), and those lacking delivery information (n=15) were excluded. A total of 465 eligible participants remained for analysis (**Figure 1**). Baseline characteristics of the participants are listed in **Table 1**. Of the total 465 participants, 11.6% (n = 54) were PE cases. 119 of 465 participants (25.6%) were with hypertensive disorders, in which 42.0% (n = 50) were PE cases.

**Figure 1.**
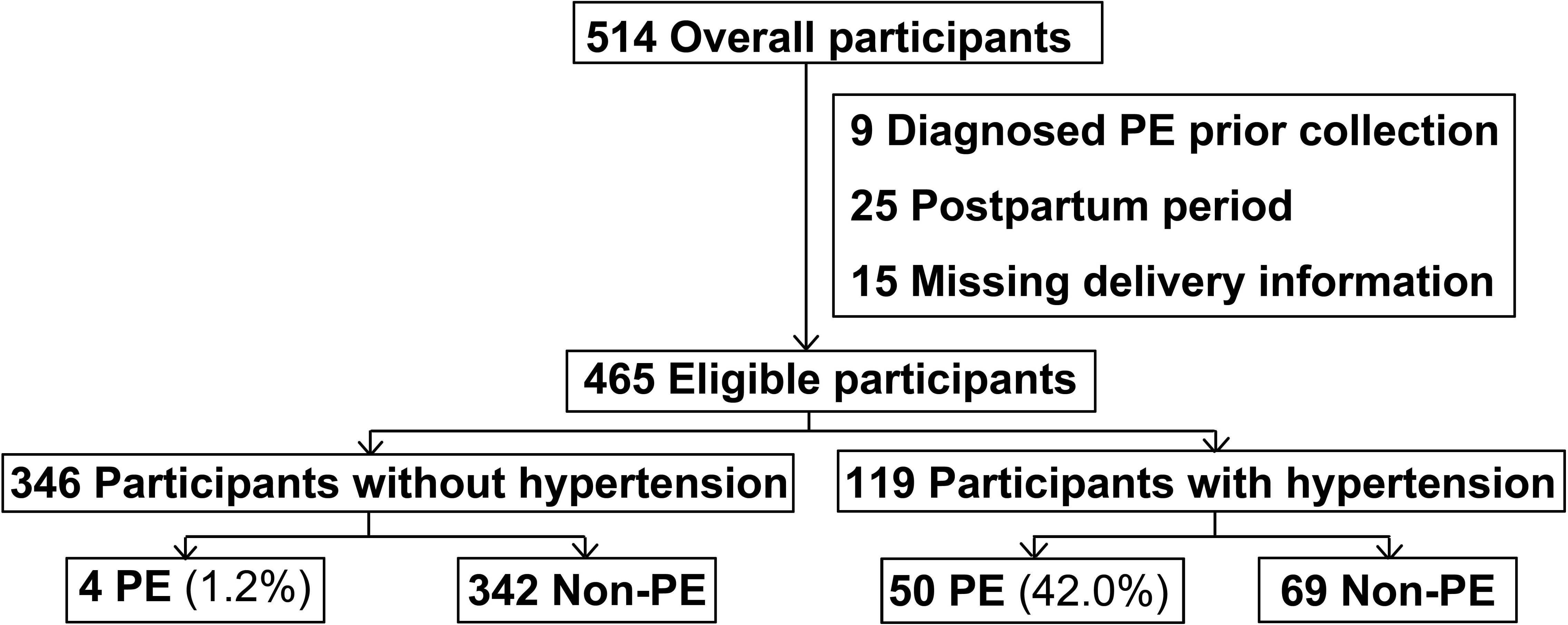
Flow diagram of participants. Hypertensive status in pregnancies was determined at the time of blood draw. (Note: Preeclampsia, PE)

**Table 1.** Baseline characteristics of the study participants.

| Characteristic | Overall (n=465) |  |  | Hypertensive (n=119) |  |  |
| --- | --- | --- | --- | --- | --- | --- |
|  | PE (n=54) | Non-PE (n=411) | *p* value<sup>^</sup> | PE (n=50) | Non-PE (n=69) | *p* value |
| Age, median (IQR), y | 35 (33-38) | 34 (31-38) | **0.040** | 35 (33-38) | 34 (32-38) | 0.151 |
|  | Race, n (%)<sup>*</sup> |  |  |  |  |  |
| *White* | 25 (46.3) | 248 (60.3) | - | 23 (46.0) | 46 (66.7) | - |
| *Asian* | 6 (11.1) | 33 (8.0) | - | 6 (12.0) | 4 (5.8) | - |
| *Black or African American* | 10 (18.5) | 35 (8.5) | **0.019** | 10 (20.0) | 7 (10.1) | 0.129 |
| *Other combinations not described* | 7 (13.0) | 54 (13.1) | - | 5 (10.0) | 7 (10.1) | - |
| *Declined* | 6 (11.1) | 41 (10.0) | - | 6 (12.0) | 5 (7.2) | - |
| GA at enrollment, median (IQR), week | 35.4 (34.0-37.0) | 29.7 (26.1-37.4) | **<0.001** | 35.6 (34.4-37.2) | 38.0 (32.9-39.1) | **0.015** |
| GA at delivery, median (IQR), week | 36.6 (35.3-37.6) | 39.0 (37.7-39.7) | **<0.001** | 36.6 (35.3-37.6) | 38.4 (37.6-39.3) | **<0.001** |
| Preterm delivery rate, n (%) | 31 (57.4) | 50 (12.2) | **<0.001** | 29 (58.0) | 8 (11.6) | **<0.001** |
| First-trimester BMI, median (IQR), kg/m<sup>2</sup> | 25.8 (22.5-30.3) | 24.6 (22.0-29.1) | 0.197 | 25.6 (22.4-30.0) | 25.5 (21.0-30.3) | 0.839 |
| C-Section, n (%) | 36 (66.7%) | 170 (41.4%) | **<0.001** | 33 (66.0%) | 35 (50.7%) | 0.140 |
| Baby weight, median (IQR), g | 2445 (2075-3105) | 3250 (2920-3540) | **<0.001** | 2550 (2133-3155) | 3175 (2655-3410) | **<0.001** |
| Nulliparous, n (%) | 38 (70.4) | 170 (41.4) | **<0.001** | 35 (70.0) | 35 (50.7) | **0.035** |
| Smoker, n (%) | 5 (9.3) | 56 (13.6) | 0.372 | 5 (10.0) | 4 (5.8) | 0.392 |
| *Current* | 2 (3.7) | 8 (1.9) | - | 2 (4.0) | 1 (1.4) | - |
| *Past* | 3 (5.6) | 48 (11.7) | - | 3 (6.0) | 3 (4.3) | - |
| *Unknown* | 0 (0) | 10 (2.4) | - | 0 (0) | 3 (4.3) | - |

Note: Preeclampsia, PE; Interquartile range, IQR; Gestational age, GA; Body-mass index, BMI.
\*: "Race" was self-reported; <sup>^</sup>The significances were calculated by Mann-Whitney U test or Chi-square test between PE and non-PE.

The median age of the PE and non-PE groups is 35 (Interquartile range, IQR, 33-38) and 34 (IQR, 31-38) respectively, showing a significant difference (*p* = 0.040) in the overall study participants, whereas there is no significant difference in age in the hypertensive subgroup. The rate of Black or African American race was significantly higher (*p* = 0.019) in the PE group (18.5%) compared to the non-PE group (8.5%), whereas there is no such significance in the hypertensive subgroup (*p* = 0.129). Conversely, the first trimester BMI and the smoking rates were not significantly different between the PE and the non-PE groups in both overall participants and the hypertensive subgroup. In contrast, the C-section rate was significantly higher in the PE group compared to the non-PE group (66.7% vs. 41.4%, *p* < 0.001) in the overall cohort; However, it was not significant in the hypertensive subgroup (66.0% vs. 50.7%, *p* = 0.140). Moreover, the baby birth weight was significantly lower in the PE group (2445 g [IQR, 2075–3105]) than in the non-PE group (3250 g [IQR, 2920–3540]), *p* < 0.001, and this difference remained significant in the hypertensive subgroup (2550 g [IQR, 2133–3155] vs. 3175 g [IQR, 2655–3410], *p* = 0.011). Nulliparous rates of the PE group are significantly higher than those of the non-PE group in both the overall participants (*p* < 0.001) and the hypertensive subgroup (*p* = 0.035). The median GA at enrollment of the PE group is significantly later than that of the non-PE group in both the overall study participants (*p* < 0.001) and the hypertensive group (*p* < 0.001). Furthermore, the median GA at delivery was 36.6 weeks for the PE group and 39.0 weeks for the non-PE group among all participants (*p*<0.001). In the hypertensive subgroup, these values were 36.6 weeks for the PE group and 38.4 weeks for the non-PE group (*p*<0.001). Deliveries occurred significantly earlier in the PE groups, and the rates of preterm delivery were significantly higher compared to the non-PE groups in both the overall study participants (*p* < 0.001) and the hypertensive subgroup (*p* < 0.001).

### sFlt-1/PlGF ratio

The baseline sFlt-1/PlGF ratio is shown in **Figure 2 (A)**. The median values of the sFlt-1/PlGF ratio for the overall cohort, the non-hypertensive subgroup, and the hypertensive subgroup are 5.5 (IQR, 2.3-29.7), 4.1 (IQR, 2.0-10.7) and 41.8 (IQR, 12.5-94.5), respectively. The baseline sFlt-1/PlGF ratio is significantly different between the non-hypertensive and the hypertensive subgroups (*p* < 0.001).

**Figure 2.**
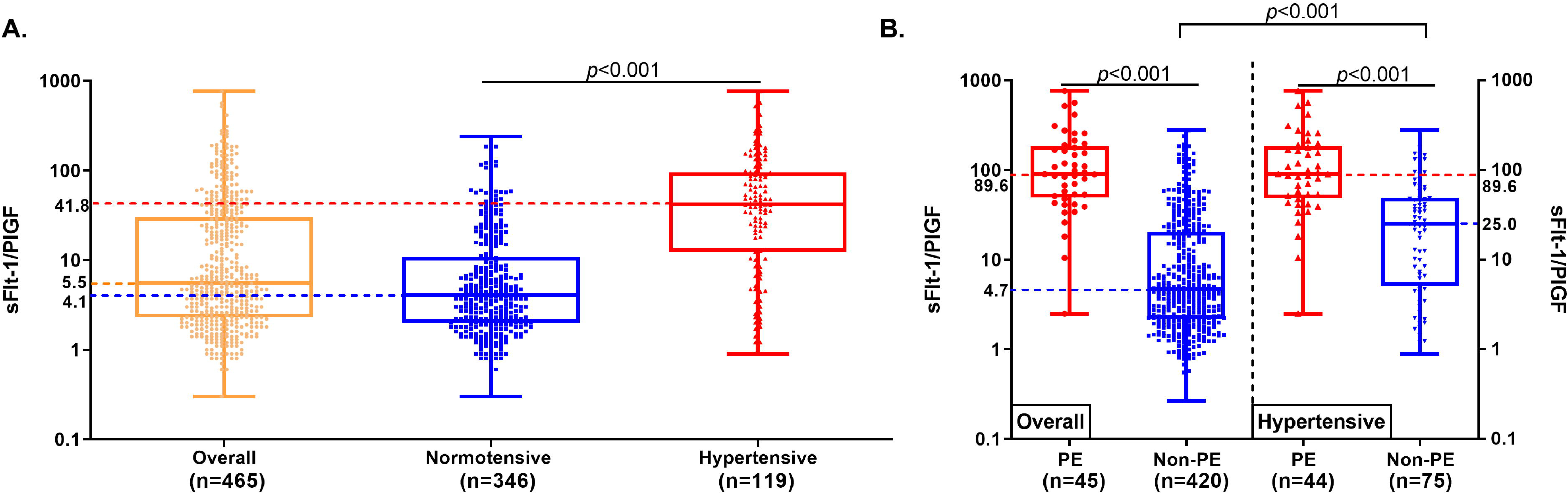
The sFlt-1/PlGF ratios in the overall participants, the non-hypertensive, and the hypertensive subgroups (**A**). The sFlt-1/PlGF ratio for participants developed preeclampsia (PE) within two weeks and those who did not (**B**). Box depicts the first (Q1) to third (Q3) quartile values, and the whiskers represent the minimum and maximum values. The *p* values were calculated by Mann-Whitney U test.

In the overall cohort, the median sFlt-1/PlGF ratio among participants who developed PE and those who did not develop PE within two weeks were 89.6 (IQR, 51.4-177.3) and 4.7 (IQR, 2.2-20.1), respectively. In contrast, the median sFlt-1/PlGF ratio among hypertensive pregnancies who developed PE and those who did not develop PE within two weeks were 89.6 (IQR, 50.4-180.3) and 25.0 (IQR, 5.7-48.5), respectively. The sFlt-1/PlGF ratio was significantly higher (*p* < 0.001) in participants who developed PE within two weeks compared to those who did not, in both the overall cohort and the hypertensive subgroup. Moreover, among participants who did not develop PE within two weeks, the sFlt-1/PlGF ratio was significantly higher (*p* < 0.001) in the hypertensive group than in the overall cohort. The results are shown in **Figure 2B**.

### ROC curve analysis and cutoff determination

The clinical performance of the sFlt-1/PlGF ratio for short-term prediction of PE within two weeks is summarized in **Table 2**. In the overall cohort (n = 465), the AUC was 0.92 (95% CI, 0.88-0.95), which was higher compared to the hypertensive subgroup (n = 119), where the AUC was 0.82 (95% CI, 0.73-0.89). The ROC curves for the overall cohort and hypertensive subgroup were shown in **Figure 3**. The optimal cutoff value for predicting PE was lower in the overall cohort (33; 95% CI, 26-47) than that in the hypertensive subgroup (50; 95% CI, 26-84). The NPV was high in both the overall cohort and the hypertensive subgroup, at 99.2% (95% CI, 98.0-100.0) and 87.6% (95% CI, 76.6-97.7), respectively. However, the PPV was higher in the hypertensive subgroup at 65.2% (95% CI, 50.0-82.5) compared to 35.3% (95% CI, 25.1-45.5) in the overall cohort. The sensitivity of the sFlt-1/PlGF ratio was 93.8% (95% CI, 85.4-100.0) in the overall cohort and 81.7% (95% CI, 61.8-97.4) in the hypertensive subgroup. Similarly, specificity was 81.3% (95% CI, 76.8-85.8) in the overall cohort and 73.1% (95% CI, 53.3-90.4) in the hypertensive subgroup. The F1-score was higher in the hypertensive subgroup (0.72; 95% CI, 0.62-0.81) compared to the overall group (0.53; 95% CI, 0.41-0.65). The LR+ was 5.74 (95% CI, 3.94-8.47) in the overall cohort and 3.49 (95% CI, 1.96-7.35) in the hypertensive subgroup, while the LR− was 0.10 (95% CI, 0.00-0.20) and 0.24 (95% CI, 0.04-0.46), respectively.

**Figure 3.**
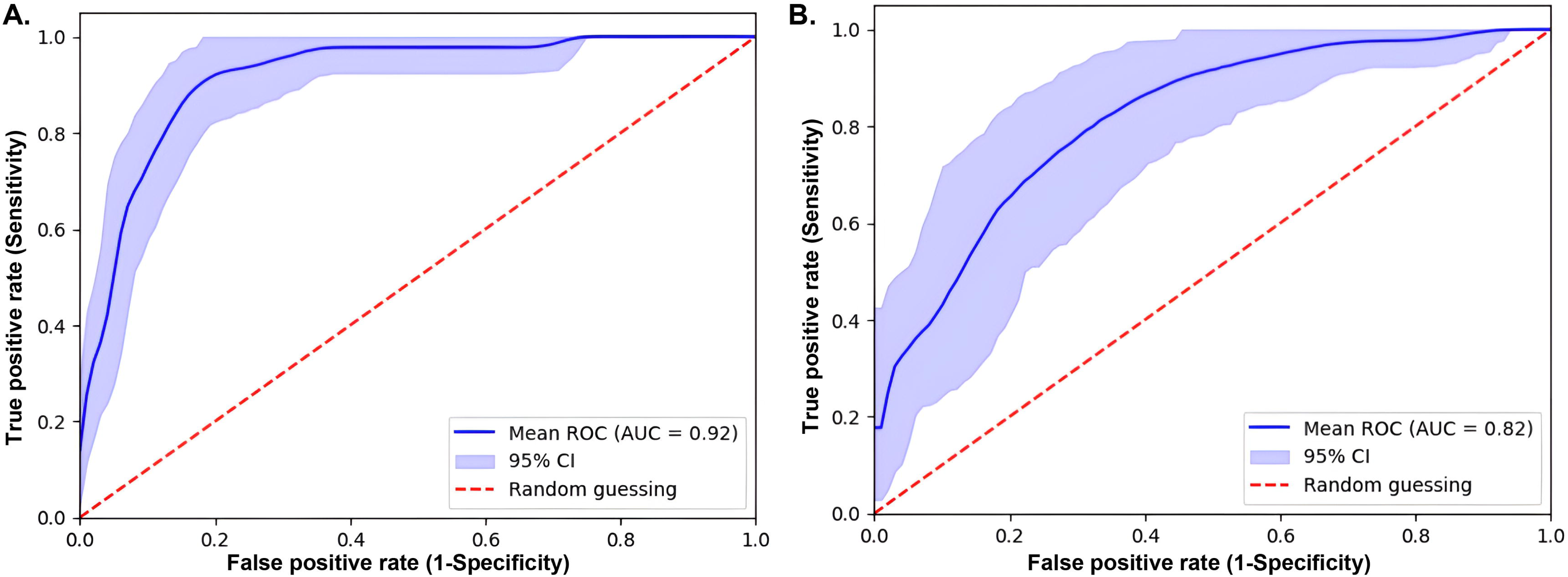
Receiver operating characteristic (ROC) curves for the overall participants(**A**) and the hypertensive subgroup (**B**).

**Table 2.** Clinical performance of the sFlt-1/PlGF ratios for short-term prediction of preeclampsia (PE) within two weeks.

| **Parameters**<br>**(95% CI)** | **Overall (n=465)** | **Hypertensive (n=119)** |
| --- | --- | --- |
| **AUC** | 0.92 (0.88-0.95) | 0.82 (0.73-0.89) |
| **Optimal cutoff** | 33 (26-47) | 50 (26-84) |
| **NPV** | 99.2% (98.0-100.0) | 87.6% (76.6-97.7) |
| **PPV** | 35.3% (25.1-45.5) | 65.2% (50.0-82.5) |
| **Sensitivity** | 93.8% (85.4-100.0) | 81.7% (61.8-97.4) |
| **Specificity** | 81.3% (76.8-85.8) | 73.1% (53.3-90.4) |
| **F1-score** | 0.53 (0.41-0.65) | 0.72 (0.62-0.81) |
| **LR+** | 5.74 (3.94-8.47) | 3.49 (1.96-7.35) |
| **LR-** | 0.10 (0.00-0.20) | 0.24 (0.04-0.46) |

**Note:** CI: Confidence interval; AUC: Area under the curve; LR+: Positive likelihood ratio; LR-: Negative likelihood ratio; NPV: Negative predictive value; PPV: Positive predictive value.

In the hypertensive subgroup, we compared the predictive performance of two different cutoffs (33 and 50) (**Supplemental Table S1**). For the cutoff of 33, the NPV was 91.3% (95% CI: 83.2-99.4), PPV was 54.8% (95% CI: 43.4-66.2), sensitivity was 90.9% (95% CI: 82.4-99.4), and specificity was 56.0% (95% CI: 44.8-67.2). In comparison, the cutoff of 50 showed comparable NPV, PPV and sensitivity but a significantly higher specificity of 73.1% (95% CI: 53.3-90.4) (*p* < 0.01).

## Discussion

This study established cutoff values and evaluated the sFlt-1/PlGF ratio’s performance in predicting PE within two weeks in a diverse U.S. pregnant population with GA ≥ 20 weeks. While the ratio effectively rules-out PE (high NPVs), its ability to rule-in PE is limited (low/moderate PPVs) and therefore the sFlt-1/PlGF ratio should be interpreted and used with cautions. Importantly, our findings highlight the need for customized cutoffs in different clinical settings and scenarios. Specifically, we demonstrated differences of the sFlt-1/PlGF ratio in the overall and hypertensive patients and observed a trend toward a higher optimal cutoff in the hypertensive subgroup compared to that in the overall study population (50 vs. 33).

Different cutoffs of sFlt-1/PlGF ratio have been reported and proposed by different studies and clinical guidelines [21,28,39,40]. The present study identified an optimal cutoff of 33 for the overall study population to rule-out PE within two weeks. Our findings align with previous studies, with 33 being consistent for ruling-out PE in a mixed population [20]. For example, Verlohren et al. also established the cutoff of 33 to aid in ruling-out PE [20] with high sensitivity (≥ 90%) in a mixed population including both non-hypertensive and hypertensive pregnant women at GA ≥ 20 weeks, which was similar to our study population. Additionally, a higher optimal cutoff of 50 was identified for hypertensive patients, indicating the cutoff should be tailored to patients with different clinical presentations. This is further supported by the comparison between the cutoffs of 33 and 50 in the hypertensive subgroup, where the higher cutoff significantly increased specificity without a notable reduction in sensitivity (**Supplemental Table S1**). The variation in optimal cutoffs between different patient groups can be attributed to varying baseline levels of sFlt-1/PlGF ratios. This study observed a significantly higher baseline of the sFlt-1/PlGF ratio in hypertensive patients compared to the non-hypertensive patients (**Figure 2**). Similarly, Verlorhen et al. reported that the sFlt-1/PlGF ratio is significantly higher (*p* < 0.001) in patients with gestational and chronic hypertension, compared to non-hypertensive pregnant patients after 34 weeks of gestation [41]. Therefore, the complex PE pathophysiology necessitates establishing customized cutoffs tailored to specific patient groups which might thus improve clinical diagnostics and management. Moreover, the PROGNOSIS study utilized a cutoff value of 38 for predicting PE within two weeks, demonstrating clinical performance comparable to that of our overall study population (**Supplemental Table S2**) [42]. Although significant differences were observed in our hypertensive subgroup, this might be explained by the PE prevalence in the PROGNOSIS study (7.5%) being more closely aligned with our overall population (9.7%) rather than our hypertensive subgroup (37.0%).

One limitation of the sFlt-1/PlGF ratio in clinical applications is the lack of a standardized cutoff, which varies among different study populations [1,28]. Further research is necessary to standardize cutoff applications across diverse populations. Utilizing the multiples of the median (MoM) method might help normalize values across different study groups, reducing variability due to factors such as race and GA [43]. Although applying different cutoffs to distinct populations might initially seem complex, expressing results as MoMs would considerably streamline interpretation. This approach would allow clinicians to consistently interpret risk, independent of underlying population characteristics or assay differences. The MoM approach standardizes results by expressing individual values relative to population-specific medians, adjusted for confounding variables. This method has been successfully applied in prenatal screening programs, such as aneuploidy screening, facilitating standardization across various populations and assay methods [44,45].

The incidence of PE varies across different population settings, and a lower PPV is typically observed in populations with a lower prevalence of PE [46]. Dragan et al. demonstrated this with a PPV of 8.3% observed for the sFlt-1/PlGF ratio at a cutoff of 38 in a routine pregnancy care population with a PE prevalence of just 1.84% [46]. In the present study, the prevalences of PE were 11.6% and 42.0% in the overall cohort and the hypertensive subgroup. The overall prevalence of PE observed in this study population is substantially higher than the 2-7% of pregnancies [2], which is not surprising as the study enrolled women with both from routine prenatal care and triage visits. Patients from triage visit usually with signs and symptoms of PE and hence at higher risk of developing PE. The prevalence of PE in the hypertensive group (42.0%) is comparable to the PRAECIS study, where the prevalence of severe PE was 33.5% among patients with hypertensive disorders [28]. The higher prevalence in our study is reasonable, as it includes both mild and severe cases of PE, where the PRAECIS study focused only on severe cases. Although the false positive rates (1-specificity) were 18.7% in the overall population and 26.9% in the hypertensive group, suggesting that approximately 20-30% of patients could receive a false alarm, the NPV values remained consistently high in both the overall population (99.2%) and the hypertensive subgroup (87.6%). These high NPVs highlight the sFlt-1/PlGF ratio’s strong capability to rule out the development of PE within two weeks. Some patients with low ratio could be safely managed as outpatient instead of inpatient. Consequently, this holds promise for reducing unnecessary admissions and alleviating the burden on healthcare resources, particularly in areas with limited medical resources. However, the PPV showed variability with a notably higher PPV in the hypertensive subgroup (65.2% vs. 35.3% in the overall population). This difference is expected due to the higher PE incidence in hypertensive subgroup (42.0%). This finding indicates that while the biomarker is better at identifying true cases of PE in hypertensive individuals, it may have limited rule-in ability in the general population. The relatively low PPV may limit clinical applications of the sFlt-1/PlGF ratio, as a high PPV is particularly important for diagnostic markers, particularly in low-prevalence conditions, such as PE [47]. Additionally, the F1-score, which balances sensitivity and PPV, was higher in the hypertensive subgroup (0.72 vs. 0.53), indicating better predictive performance in predicting true positives in hypertensive patients.

Although the sFlt-1/PlGF ratio has shown promising performance in predicting PE, limitations such as low PPV, variable cutoff values, and lack of standardization remain. Recent advances in artificial intelligence and machine learning have enabled the development of models that integrate clinical risk factors and additional biomarkers to predict the onset of PE. These AI-based models—incorporating factors such as BMI, age, parity, and preexisting medical conditions—have demonstrated very promising performance [48,49]. However, few studies have included the sFlt-1/PlGF ratio in these models, likely due to its limited clinical availability. Future research may focus on developing AI-based models that incorporate the sFlt-1/PlGF ratio to further enhance the prediction of PE and improve maternal and fetal outcomes.

Our study has several limitations. First, the cutoff values are derived from Roche’s assays, and there may be variability using other assays. Second, despite the diversity of our population, this was a single-center study, and external validation is needed to reduce the risk of bias. Third, due to low PE prevalence and insufficient sample size, we couldn’t perform cutoff analysis in the non-hypertensive pregnant patient subgroup, limiting the generalizability of our findings. Fourth, the significant difference in GA at the time of blood draw between the PE and non-PE groups could potentially influence diagnostic performance, especially considering that the sFlt-1/PlGF ratio naturally rises in non-hypertensive women after 28 weeks of gestation [50]. Fifth, given a relatively small size in the hypertensive subgroup, a simple hold-out validation approach can introduce substantial bias and high variability, where different randomizations can yield different optimal cutoff values. It explains our previously reported optimal cutoff of 38 in our preliminary study using a simple hold-out validation approach [51]. To mitigate the potential bias associated with this approach, we employed cross-validation with bootstrapping – a robust statistical technique that provides a more stable and reliable estimate of model performance by averaging results over multiple splits [52,53]. Specifically, the optimal cutoff of 50 with a wide range of 95%CI (26-84) was identified in the hypertensive patients using stratified 5-fold cross-validation with bootstrapping. However, to achieve a narrower confidence interval range, a larger sample size is needed to validate our results and better understand the sFlt-1/PlGF ratio’s performance in a broader context.

## Conclusions

Our study evaluated the clinical performance of the sFlt-1/PlGF ratio in predicting PE within two weeks in a diverse U.S. population. It suggests that while the sFlt-1/PlGF ratio is highly effective for ruling-out PE, its ability to rule-in is more limited. Additionally, our findings suggest that there is a need for customized cutoffs in different clinical settings, and a higher sFlt-1/PlGF ratio has been observed in the hypertensive patients therefore a higher cutoff is likely essential for accurately ruling-in/out PE in these patients. To optimize the clinical use of the sFlt-1/PlGF ratio, further research is needed to establish standardized cutoffs, evaluate performance across different assays, and explore broader clinical applications, such as in outpatient settings. Tailoring the sFlt-1/PlGF ratio to specific patient populations and clinical contexts can potentially enhance its effectiveness in predicting and managing PE.

## Supplemental Material

See the attached word files.

## Acknowledgments

We would like to thank all patients participating in this study.

## Author contributions statement

Yaxin Li (Date curation, formal analysis, investigation, writing-original draft and writing-review & editing), Kristen Cagino (Date curation, investigation, methodology and writing-review & editing), Jim Yee (Data curation and formal analysis), Caroline Andy (Formal analysis), Dajana Borova (Conceptualization and project administration), Ayush Shah (Data curation and formal analysis), Isla Racine (Formal analysis and writing-review & editing), Tracy Grossman (Conceptualization, data curation, investigation, methodology, project administration and writing-review & editing), Zhen Zhao (Conceptualization, investigation, project administration, resources, supervision and writing-review & editing). All authors have read and approved the final version of the manuscript.

## Data availability statement

The datasets used and analyzed in this study are available from the corresponding author upon reasonable request.

## Additional information

## Nonstandard Abbreviations

PE: Preeclampsia
U.S.: the United States
sFlt-1: Soluble fms-like tyrosine kinase-1
PlGF: Placental growth factor
GA: Gestational age
NICE: National Institute for Health and Care Excellence
IRB: Institutional review board
BMI: Body mass index
ACOG: American College of Obstetricians and Gynecologists
EMR: Electronic medical records
PPV: Positive predictive value
NPV: Negative predictive value
CI: Confidence interval
ROC: Receiver operating characteristic
AUC: Area under the curve
IQR: Interquartile range
LR+: Positive likelihood ratio
LR-: Negative likelihood ratio
MoM: Multiples of the Median

## Research Funding

This study is sponsored by Roche Diagnostics and Clinical and Translational Science Center (CTSC) at Weill Cornell Medicine (Grant No. UL1TR002384).

