## Supplemental materials for "Short-term Preeclampsia Prediction: Cutoff Variations for sFlt-1/PlGF in U.S. Patients with or without Hypertensive Disorders"

**Table S1.** Comparative analysis using two different cutoffs 33 and 50 in hypertensive group (n=119).

| **Parameters**  **(95% CI)** | **Cutoff 33** | **Cutoff 50** |
| --- | --- | --- |
| **NPV** | 91.3% (83.2-99.4) | 87.6% (76.6-97.7) |
| **PPV** | 54.8% (43.4-66.2) | 65.2% (50.0-82.5) |
| **Sensitivity** | 90.9% (82.4-99.4) | 81.7% (61.8-97.4) |
| **Specificity** | 56.0% (44.8-67.2) | 73.1% (53.3-90.4) ** |

**Note:** The cutoff of 33 and 50 were established in the current study using the stratified 5-fold cross-validation with bootstrapping method in the overall study cohort and hypertensive cohort, respectively. ***p*<0.01 represents specificity significantly higher for the cutoff 50 compared to the cutoff 33.

**Table S2.** Comparison analysis for the sFlt-1/PlGF ratio in predicting preeclampsia (PE) within two weeks.

|  | **Overall**  **(n=465)** | **Hypertensive pregnancies (n=119)** | **PROGNOSIS study**  **(n=550)** |
| --- | --- | --- | --- |
| **Cutoff** | 38 | | |
| **NPV (95% CI)** | 98.4% (97.1-99.7) | 88.7% (80.1-97.2) ******* | 97.9% (96.0-99.0) |
| **PPV (95% CI)** | 39.4% (29.8-49.0) | 57.6% (45.7-69.5)  ****** | 32.0% (22.9-41.1) |
| **Sensitivity (95% CI)** | 86.7% (76.7-96.6) | 86.4% (76.2-96.5) | 78.0% (62.4-89.4) |
| **Specificity (95% CI)** | 85.7% (82.4-89.1) | 62.7% (51.7-73.6) ******* | 81.1% (77.5-84.4) |

**Note:** “CI” stands for Confidence interval. A Z-test was applied to compare different groups in current study with PROGNOSIS study (Validation cohort). The significance levels were denoted as follows: ***p* < 0.01, ****p* < 0.001.
